# Development of a Target Product Profile for a Proteomic Blood Test to Aid the Diagnosis of Cranial Giant Cell Arteritis - *Study protocol*

**DOI:** 10.64898/2026.09.07.26362451

**Authors:** Ann W. Morgan, Sarah L. Mackie, Mark M. Iles, Miaoqing Yang, Paola Cocco, Amy Rebane, James I. Robinson, Louise Sorensen, Michael P. Messenger

## Abstract

**Background:** Giant cell arteritis (GCA) is the commonest systemic vasculitis and a medical emergency. High-dose glucocorticoids are started on suspicion, before confirmation, and cause substantial dose-dependent harm in the many patients who prove not to have GCA. Diagnostic confirmation by temporal artery biopsy or ultrasound remains imperfect. The performance of both approaches is influenced by the clinical context and, for ultrasound, operator expertise, creating the potential for both unnecessary treatment and delayed or missed diagnosis. A laboratory-based, multi-analyte proteomic blood test may help, but its intended use and performance requirements are undefined.

**Objective:** To develop, by structured expert consensus, a Target Product Profile (TPP) for a laboratory-based, multi-analyte proteomic blood test to aid the diagnosis of cranial GCA in secondary care, used alongside clinical assessment, imaging and/or biopsy, with the primary objective of reducing unnecessary glucocorticoid exposure without increasing missed cases, from a UK health-system perspective.

**Methods and analysis:** A multidisciplinary committee of approximately 15 to 30 members, including patient and public representatives, will follow a three-phase process: a scoping phase that confirms the intended use and the prioritised list of characteristics; a drafting phase that specifies minimum (acceptable) and preferred (desirable) values across the TPP domains, using outcome-based methods where feasible; and a consensus-building phase using an up-to-three-round modified Delphi (75% consensus threshold) with interim consensus meetings and a 28-day public consultation. Reporting follows ACCORD and CREDES.

## 1. Background and rationale

Giant cell arteritis (GCA) is the commonest primary systemic vasculitis and a medical emergency: untreated cranial disease can cause irreversible sight loss within days. Because of this risk, high-dose glucocorticoids are started on clinical suspicion, before diagnosis is confirmed^1^. Confirmatory diagnostic testing is imperfect but recommended for all patients with suspected GCA^1, 2^. Temporal artery biopsy is highly specific but has limited sensitivity, while ultrasound is more sensitive, less specific and is operator-dependent^3^. As glucocorticoid treatment can reduce imaging signs of vascular inflammation, ultrasound should be performed within 72 hours of glucocorticoid initiation^2^. False-positive diagnoses may expose patients without GCA to prolonged glucocorticoid treatment and associated adverse effects^1^. Whilst delayed or missed diagnosis increases the risk of irreversible ischaemic complications^4^.

Plasma proteomic profiling has identified an 11-protein signature that distinguishes biopsy-proven cranial GCA from its clinical mimics with high discrimination (AUC 0.94)^5^. Early cost-utility modelling from a UK NHS perspective indicates that a sufficiently accurate blood test could be cost-effective, with value driven more strongly by specificity than by sensitivity^6^. This document outlines the protocol for development of a Target Product Profile (TPP), through a structured multidisciplinary expert consensus process, to agree upon the minimum (acceptable) and preferred (desirable) characteristics such a test must meet to be fit for purpose^7–9^.

## 2. Aim and objectives

### Aim

To establish, through structured expert consensus, a Target Product Profile for a *laboratory-based, multi-analyte proteomic blood test to aid the diagnosis of cranial GCA in secondary care, used alongside clinical assessment, imaging and/or biopsy*, with the primary objective of reducing unnecessary glucocorticoid exposure without increasing missed cases, from a UK health-system perspective.

Objectives, mapped to the three development phases:

1. **Scoping phase:** confirm and refine the intended use, including evaluating whether a direct-to-test positioning is feasible and preferable and should form the dedicated scope of the TPP, and agree the prioritised list of characteristics to be specified.
2. **Drafting phase:** define the candidate characteristics and draft, for each, the minimum (acceptable) and preferred (desirable) values.
3. **Consensus-building phase:** reach consensus on the specifications through an iterative Delphi process and open public consultation, and publish the final agreed TPP with its consensus results.

## 3. Draft Scope

The following draft scope will be reviewed by the TPP committee and, where necessary, refined during the scoping phase (Section 5.2).

| Parameter | Proposed scope |
| --- | --- |
| Technology | Laboratory-based, multi-analyte proteomic blood test. |
| Intended use | To aid the diagnosis of cranial GCA in secondary care, used alongside clinical assessment, imaging and/or biopsy (adjunctive |
|  | diagnostic aids, not stand-alone replacements for clinical judgement). |
| **Primary objective** | Reduce unnecessary glucocorticoid exposure without increasing missed cases resulting in major ischaemic events including sight loss. |
| **Target condition** | Cranial GCA. |
| **Target population** | Adults with suspected cranial GCA, with or without prior glucocorticoid treatment. |
| **Setting** | Secondary care (specialist clinics), with blood sampled within secondary care. |
| **Perspective** | United Kingdom health-system perspective. |
| **Out of scope** | Monitoring or relapse detection; pure large-vessel (extracranial) GCA; non-laboratory test modalities; and non-UK perspectives. |

## 4. Governance and committee

A multidisciplinary TPP Development Committee of approximately 15 to 30 members will be convened, chaired by Prof. Morgan, with a second independent co-chair to be appointed, and facilitated by an experienced TPP consultant (Prof. Messenger). Membership will span the disciplines relevant to the pathway: rheumatology, ophthalmology, primary care, clinical laboratory and biomedical science, histopathology, medical imaging, pharmacy, health economics, statistics or methodology, regulation and health policy, and patient and public representatives.^[2,9]^

Committee members will be identified through two complementary routes. First, systematic stakeholder mapping, informed by a modified 7Ps Framework^10^ (see table 1), will identify relevant UK organisations, professional groups, disciplines and patient and public perspectives. The resulting stakeholder matrix will be reviewed for gaps in representation and supplemented by snowball sampling. Second, a predefined literature search will identify academic experts, informed by relevant publication output and citation impact. Experts identified through the publication/citation analysis will comprise fewer than half of the committee, so that the panel retains broad pathway and stakeholder representation. In keeping with the UK scope of the TPP, membership will be limited to UK-based participants and all physicians will be consultants who are currently practicing in the NHS. All members will complete a declaration of interest form, reviewed by the Chairs before participation.

**Table 1.**
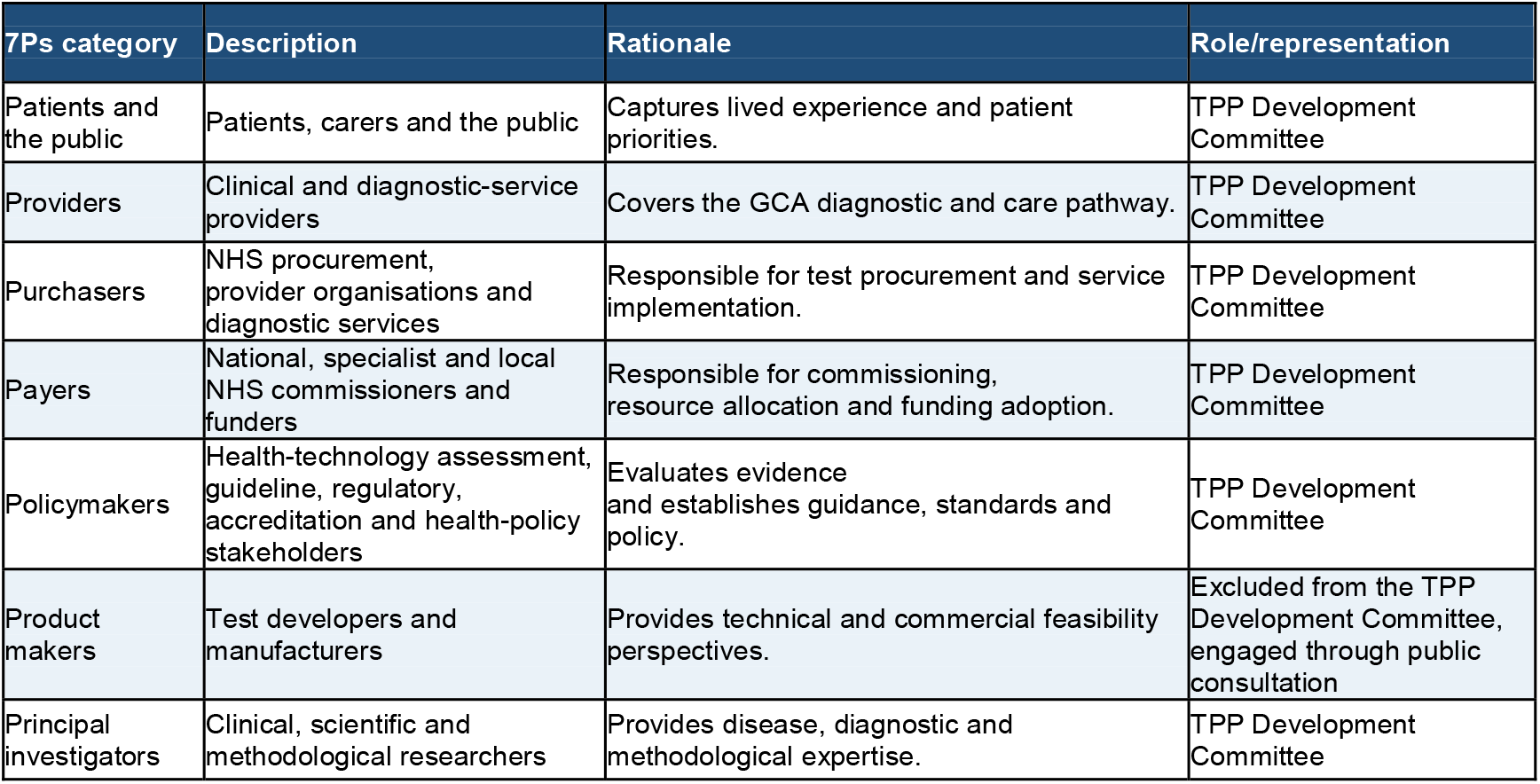
Adaptation of the 7Ps framework^10^ for the GCA TPP.

Consensus meetings will be quorate when more than half of members attend. Committee members will be expected to participate throughout the consensus process; members who do not complete a Delphi survey within 14 days, following up to two reminders, will be considered to have withdrawn from the committee and will not be invited to subsequent rounds, unless exceptional circumstances are agreed with the Chairs. Completion of the consensus process, together with meeting the other applicable authorship criteria, will normally be required for authorship of the final publication. The adequacy of each Delphi round will be assessed against the overall response rate and response rates, withdrawals and changes in panel composition between rounds will be reported.

## 5. Methodology

The TPP will be developed over three phases, scoping, drafting and consensus-building, adapted from the WHO procedure and the framework of Cocco and colleagues, and will be reported in line with ACCORD and CREDES^7–9, 11^. The scoping phase determines the intended-use decision that shapes everything that follows.

### 5.1 Phase 1: Scoping

In the scoping phase the committee will assess the proposed secondary-care use case and evaluate whether other use cases may be feasible or preferable. The committee may confirm the proposed scope, refine it, or determine that an alternative use case should become the dedicated scope of the TPP. The discussion will draw on the platform outputs above. The output will be a confirmed intended use, scope and prioritised list of characteristics, with a documented rationale, which will form the basis for drafting.

### 5.2 Phase 2: Drafting

A multidisciplinary working group will populate the TPP structure across its standard domains: clinical scope; test scope, including measurand and specimen; analytical performance; clinical performance; operational performance; human factors; infrastructure; and additional economic, regulatory and environmental considerations^7^. Draft minimum and preferred values will be prepared by the Steering Group, with the supporting evidence documented for each.

The group will seek to derive outcome-based analytical and clinical performance specifications where possible, using appropriate simulation and decision-analytic methods^12, 13^.

### 5.3 Phase 3: Consensus-building

Consensus will be developed through an up-to-three-round modified Delphi. Rounds 1 and 2 will be used to rate and refine the draft specifications; following public consultation, a final round will be limited to unresolved characteristics and those materially revised in response to consultation. In each round members will rate every minimum and preferred criterion on a 4-point Likert scale (fully disagree, mostly disagree, mostly agree, fully agree), with an explicit no-opinion option, and will be required to comment when they disagree; minimum and preferred specifications will be rated independently^8, 11, 14^.

Consensus for inclusion will be defined a priori as at least 75%^15^ of participants providing an evaluative response and selecting mostly agree or fully agree. No-opinion responses and missing responses will be excluded from the denominator but reported separately, and the full response distribution will be retained. The 75% threshold has been selected a priori as a conventional threshold and corresponds to the median percentage-agreement threshold identified by Diamond and colleagues in their systematic review of Delphi studies^15^.

Individual survey responses will remain confidential to the analysis team and will be presented to the committee only in aggregate. Consensus meetings will explore the reasons for disagreement and agree proposed revisions; consensus status will be determined through the subsequent independent Delphi rating rather than by discussion. Items not reaching consensus will be revised, excluded or recorded as unresolved.

#### Public consultation

The revised draft will be released for open public consultation for 28 days, promoted by committee members by email and social media. Consultation responses will be reviewed as advisory evidence and will not themselves constitute consensus votes. Respondents will be asked to identify their stakeholder category and relevant interests; responses from test developers and parties with a direct commercial interest will be identified and summarised separately, while remaining eligible to inform technical feasibility and development considerations. Responses will be collected through an online form, reviewed by the Chairs and facilitators, and used to revise the draft.

#### Final consensus and sign-off

Consultation responses and proposed changes will be discussed at a final consensus meeting and circulated for a final Delphi round, which will be limited to unresolved characteristics and those materially changed in response to consultation (a material change being one that affects the meaning, scope or value of a characteristic). Characteristics not reaching 75% agreement after this round will be either excluded or recorded as unresolved, with a narrative explaining the disagreement and highlighting considerations for further research.

## Data Availability

All data produced in the present work are contained in the manuscript

## 6. Ethics and patient and public involvement

This is a consensus-development study in which professional and public contributors act as co-investigators and members of the TPP Development Committee. It does not recruit patients or other individuals as research participants and does not involve patient data, biological samples or clinical interventions, and Research Ethics Committee review is therefore not required. Patient and public involvement will be embedded throughout, coordinated through the NIHR Leeds BRC PMR/GCA involvement group and national partners, and public contributors will be reimbursed in accordance with NIHR guidance.

## 7. Outputs and dissemination

This protocol will be posted as a preprint on medRxiv (https://www.medrxiv.org/). The agreed TPP, the Delphi results (percentage agreement and response counts) and a summary of consultation feedback will be compiled into a final report and prepared for peer-reviewed publication, reported in line with ACCORD and CREDES^8, 11^. The finalised characteristics will serve as a benchmark for downstream analysis of the proteomic signature and to signal requirements to test developers and funders.

## Declarations

### Author contributions

AWM conceived and leads the programme. MPM and PC designed the TPP methodology. MPM will facilitate the consensus process. MY and PC lead the health-economic analysis. SLM provides clinical and guideline expertise. MMI provides statistical and methodological input. AR leads patient and public involvement. JI provides laboratory science and proteomics expertise. LS provides project management. All authors reviewed and approved the protocol.

### Data availability

De-identified aggregate voting results, item-level response counts, revisions between rounds and the final TPP will be made publicly available. Free-text responses will be shared only in anonymised or summarised form to protect contributor confidentiality.

